# Interventional Rescue Therapy for Delayed Cerebral Ischemia after Aneurysmal Subarachnoid Hemorrhage: a 10-Year Single-Center Experience

**DOI:** 10.64898/2026.08.25.26361378

**Authors:** Cédric Kissling, Thomas Petutschnigg, Danial Nasiri, Johannes Goldberg, David Bervini, Tomas Dobrocky, Eike I. Piechowiak, Michael Murek, Mandy D. Müller, Philippe Schucht, Joerg C. Schefold, Andreas Raabe, Werner Z’Graggen

**Author notes:** Corresponding Author: Cédric Kissling, MD, Dr. med. Cédric Kissling, Universitätsklinik für Neurochirurgie, Inselspital, Rosenbühlgasse 25, 3010 Bern.

## Abstract

**Background:** Evidence regarding delayed cerebral ischemia (DCI) after aneurysmal subarachnoid hemorrhage (aSAH) remains sparse. We aimed to identify its predictors and occurrence and evaluate its role in ischemic stroke and functional outcome under treatment with interventional rescue therapy (IRT).

**Methods:** This retrospective single-center study included 628 adults with aSAH from 2014– 2023. The primary endpoint was occurrence of refractory DCI (= refractory despite induced hypertension) treated with at least one IRT. Multivariable models evaluated refractory DCI, new ischemic stroke, and poor functional outcome (mRS 3–6) at 6–12 months.

**Results:** Among 628 included patients, 61 who died within 3 days were excluded from DCI analysis; 166/567 (29%) developed refractory DCI. Younger age (OR = 0.98; *P*<0.001), female sex (OR = 0.57; *P*=0.007), and higher WFNS grade (OR = 1.18; *P*=0.011) were independently associated with refractory DCI. Earlier first IRT was associated with longer DCI duration (IRR = 0.88; *P*<0.001) and more required IRTs (IRR = 0.91; *P*<0.001). IRT was performed later than day 14 in 29/166 patients (17.5%); none was older than 70 years. Refractory DCI was associated with new ischemic stroke (OR = 4.68; *P*<0.001) and poor functional outcome (OR = 2.37; *P*<0.001); earlier first IRT was associated with poor outcome within the refractory DCI subgroup (OR = 0.86; *P*=0.03). Outcomes after 1–2 IRTs did not differ from those without refractory DCI (*P*=0.4), whereas ≥3 IRTs were associated with poor outcome (*P*=0.04).

**Conclusions:** Refractory DCI affected 29% of aSAH patients, predominantly younger women and patients with poorer initial neurological status, and extended beyond day 14 in nearly 20% of affected patients, none of whom was older than 70 years. Refractory DCI and earlier onset were associated with poorer radiological and functional outcomes. The absence of a detected outcome difference after 1–2 IRTs suggests that favorable outcomes may remain achievable despite refractory DCI.

## Introduction

Delayed cerebral ischemia (DCI) is observed in approximately 20-41% of patients after aneurysmal subarachnoid hemorrhage (aSAH) and is a major determinant of poor outcome, including death. ^1–8^ DCI is defined by new focal neurological deficits or a reduced conscious state days to weeks post-ictus not attributable to other than cerebral hypoperfusion. ^1,9^

Early DCI results from SAH-induced neuroinflammation and endothelial dysfunction, leading to cortical spreading depolarizations and hemodynamic changes, including a loss of autoregulation. ^10^

Late DCI is primarily caused by cerebral vasospasm (CVS), first described in 1951 and extensively studied since. ^10,11^ CVS is associated with blood degradation products, cortical spreading depolarizations, and cytotoxic changes in the cerebral vasculature. ^10^ DCI typically occurs 4 to 21 days after the initial aSAH, peaking around days 8 to 10 and declining gradually thereafter. ^8,12–15^ While neither occurrence nor severity of DCI can be predicted reliably so far, previous work has associated occurrence of DCI to various clinical findings, blood markers and radiological parameters. ^16–35^

Oral nimodipine is the so far only pharmacologic concept with a proven benefit by lowering the occurrence of cerebral infarctions and thus poor clinical outcome. ^36–40^ Therapy-wise, while induced hypertension is widely recommended for DCI backed up by observational data, hypervolemia and hemodilution (as further components of the former “Triple-H” therapy) have been discouraged in favor of euvolemia. ^1,5^ Still, randomized trials have failed to demonstrate clear benefits of induced hypertension regarding long-term clinical outcome. ^1,5^ In case of persisting neurological deficits despite induced hypertension, a condition referred to as ‘refractory DCI’, many centers advocate interventional rescue therapies (IRT) such as intraarterial nimodipine administration, percutaneous transluminal balloon or stent angioplasty. ^1,41–48^ However, these procedures carry significant, mostly catheter-related risks, are often short-acting and therefore may need to be repeated in case of DCI recurrence. ^43^ Moreover, the supporting evidence remains limited and the current guidelines from 2026 strongly recommend further investigation. ^49^ Other proposed forms of IRTs include continuous intraarterial and intrathecal nimodipine application, while current evidence remains preliminary. ^50–55^

We conducted a retrospective analysis assessing the occurrence and severity of refractory DCI, its association with clinical and radiological parameters and functional outcome after IRT.

## Methods

### Study Design and Ethics

We conducted a single-center, retrospective analysis at our tertiary hospital. All adult patients diagnosed with aSAH from January 2014 to December 2023 were included. Patients were excluded if procedural data or radiological information were missing, or if they refused to provide consent. To limit bias in the assessment of DCI occurrence, severity, and impact, patients who died within three days after the ictus were excluded from further analysis. This study is reported in accordance with the Strengthening the Reporting of Observational Studies in Epidemiology (STROBE) statement. ^56^ The local ethics committee approved the study (KEK Bern 2024-01289).

### Patient Management

All patients were treated according to our institutional standard of care, which includes hourly neurological examinations at the intensive or intermediate care unit for at least 12 days after initial aSAH. All patients underwent cerebral imaging (MRI or CT) the day after endovascular or surgical aneurysm treatment (within less than 24 hours) to assess treatment-related complications. In case of neurological deterioration (defined by a decrease of at least 2 points on the Glasgow Coma Scale or a new focal neurological impairment) lasting for at least one hour (not attributable to other causes and thus indicating DCI), emergency CT imaging including angiography and perfusion imaging was performed. If DCI was confirmed by detection of regional perfusion deficit in time-to-drain sequences, it was primarily managed by induced hypertension (increase of mean arterial pressure by 10-20 mmHg) and euvolemia. ^57^ If neurological deficits persisted despite induced hypertension (= ‘refractory DCI’), endovascular transarterial local nimodipine administration was used as first-line IRT. In severe cases of refractory DCI, continuous intraarterial and/or intrathecal nimodipine administration were performed.

### Primary and Secondary Endpoints

The primary endpoint was the occurrence of refractory DCI, treated by at least one performed IRT. All patients meeting this endpoint were compared to the remaining patients (with either no signs of DCI or non-refractory DCI) (see Figure 1) regarding secondary endpoints: baseline demographics *(sex, age)*, radiological data *(the aneurysm-harboring vessel, aSAH scores such as Fisher and BNI*^58^*, the occurrence of an intracerebral hemorrhage, a new ischemic stroke at discharge compared to the immediate imaging after aneurysm treatment)*, treatment details (*endovascular, microsurgical or no treatment, external ventricular and lumbar drainage*), and outcome measures (*the number of days hospitalized, deaths during hospitalization, discharge destinations and the mRS at follow-up*) (see Tables 1 and 2). Furthermore, all DCI patients were assessed regarding the occurrence (*the first and last day as well as the duration*) and the severity (*the number of required IRTs*) of refractory DCI.

**Figure 1.**
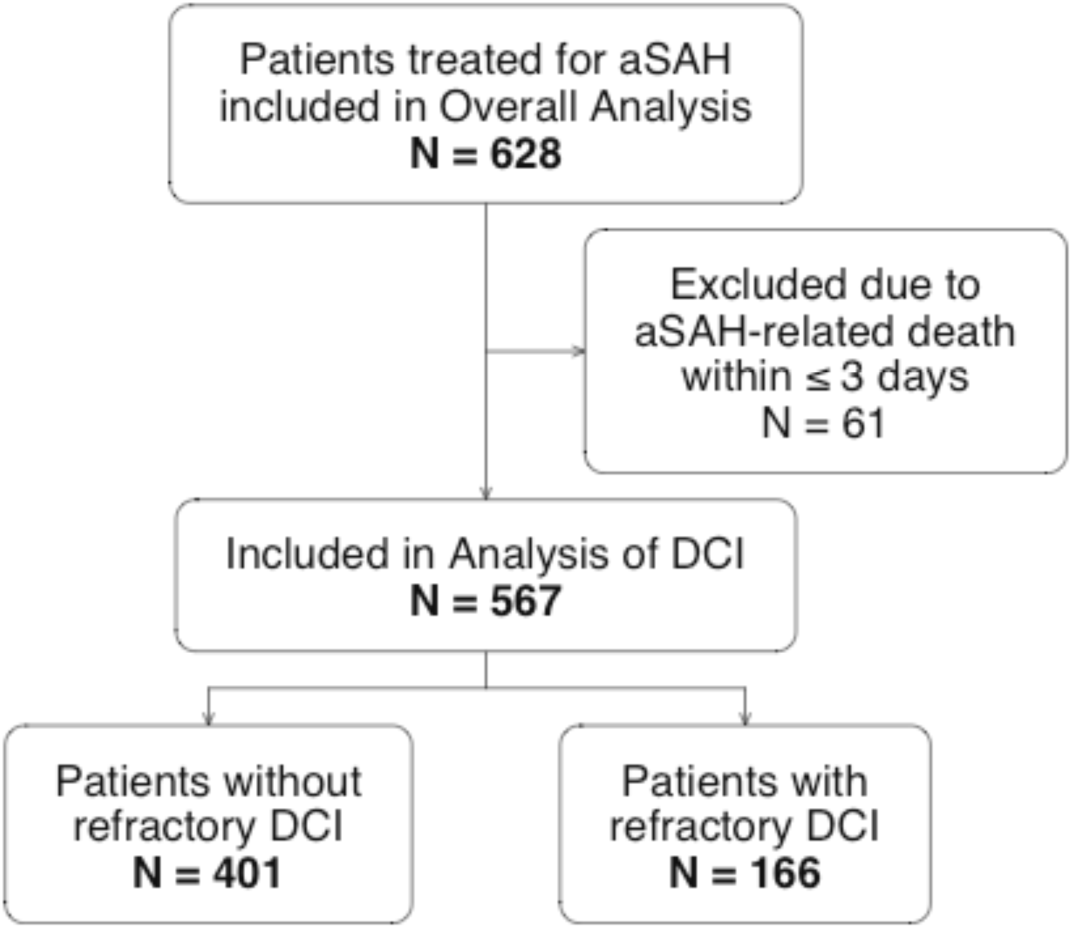
Flow Chart of Patient Inclusion. aSAH = aneurysmal Subarachnoid Hemorrhage, DCI = Delayed Cerebral Ischemia

**Table 1.**
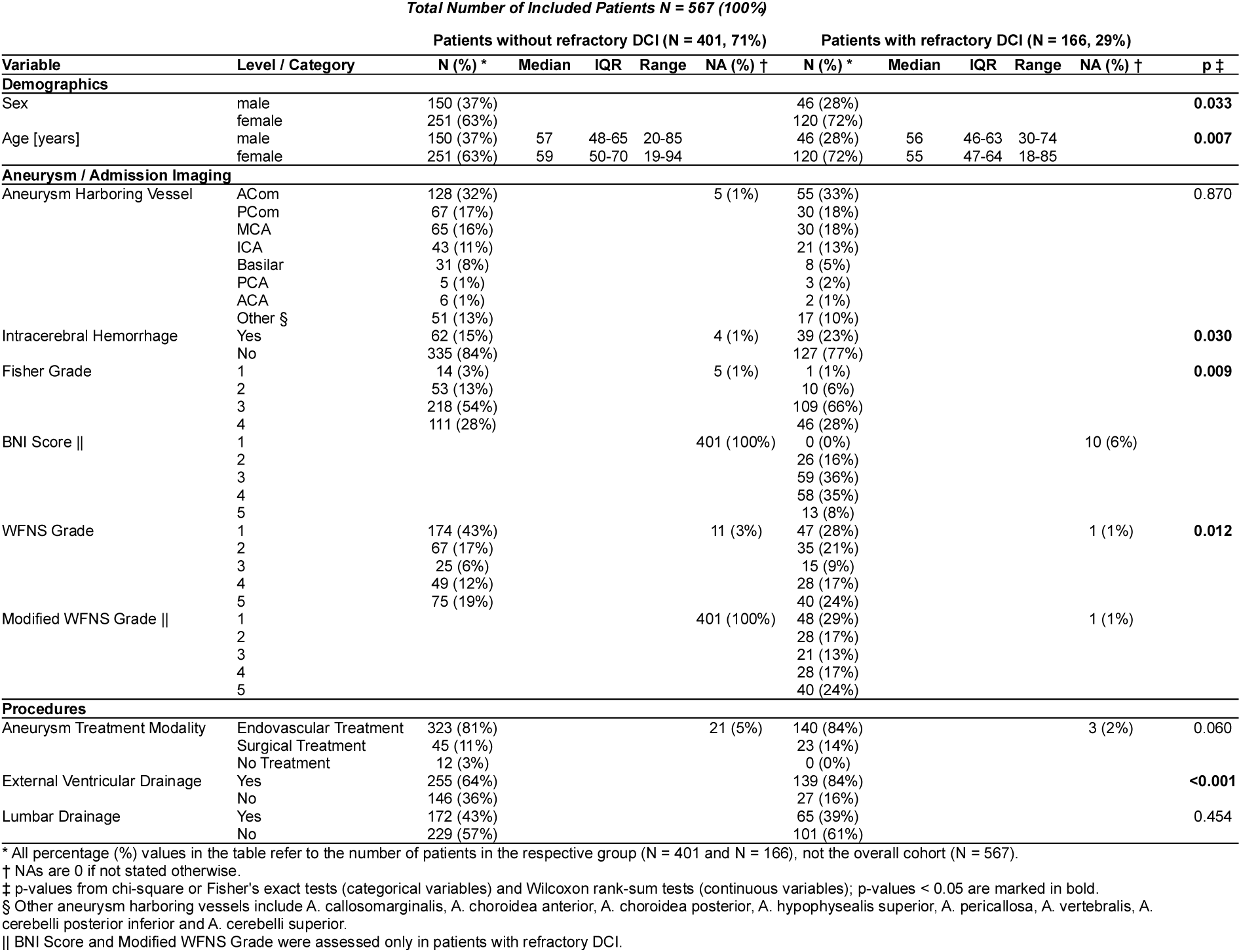
Demographics, Characteristics, and Procedures in Patients with and without refractory DCI.

**Table 2.**
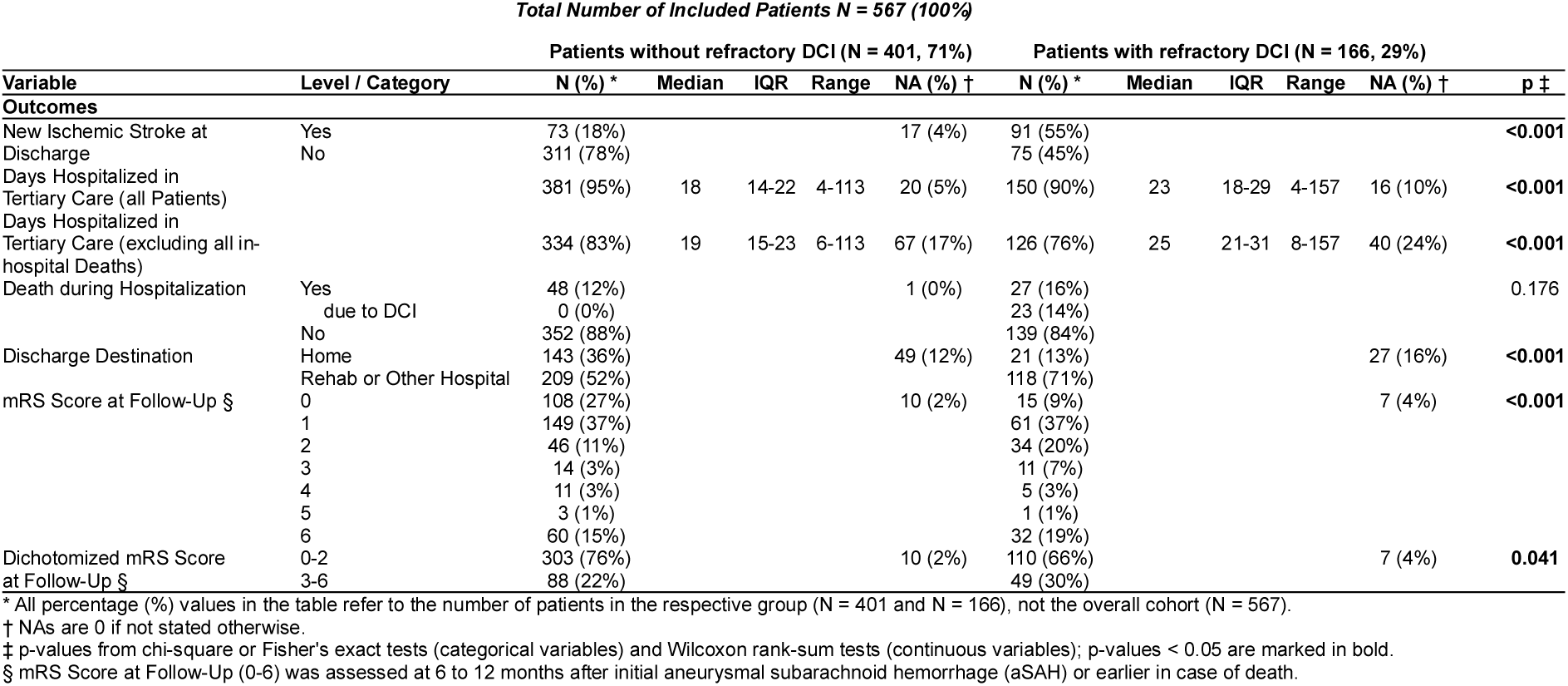
Radiological and Clinical Outcome in Patients with and without refractory DCI.

### Radiological Assessment

For radiological evaluation, Sectra Workstation IDS7 software (Version 24.2, Sectra AB, Linköping, Sweden) was used.

### Statistical Analyses

All analyses were conducted using RStudio Version 2024.09.1+394 (released 2024, Posit Software, PBC). Normality of continuous data was assessed using the Shapiro– Wilk test. Continuous variables are reported as median and interquartile ranges; categorical variables as counts and percentages. Group comparisons for categorical variables were performed using Chi-square tests (with Yates’ continuity correction) or Fisher’s exact tests, when appropriate. Comparisons of ordinal or continuous variables were performed using Wilcoxon rank-sum tests. A directed acyclic graph (DAG) was created on DAGitty Version 3.1 (updated 2023, J. Textor, Radboud University, Nijmegen, Netherlands) to assess potential bias in our empirical multivariable models. ^59,60^ Multivariable binary logistic regression models were then used to assess adjusted associations with (1) occurrence of refractory DCI, (2) new ischemic stroke at discharge, and (3) functional outcome at follow-up (dichotomized mRS 3–6 vs 0–2) in all patients surviving >3 days after initial aSAH. In the subgroup of patients treated interventionally for refractory DCI, multivariable models included linear regression for the day of first and the day of last IRT, negative binomial regression for duration of refractory DCI, Poisson regression for number of IRTs, and logistic regression for new ischemic stroke and dichotomized mRS at follow-up. As local nimodipine administration through an endovascular approach was the most common type of IRT, the “number of IRTs” included only these. Multicollinearity was assessed using Spearman correlation matrices and variance inflation factor (VIF/GVIF-equivalent) analyses. Model adequacy was assessed using dispersion measures (negative binomial θ; Poisson overdispersion φ), and goodness-of-fit metrics including AIC and pseudo-R²/adjusted R² where applicable. For a subgroup comparison of functional outcome based on the number of performed IRTs for refractory DCI, Wilcoxon rank-sum shift test for ordinal mRS and Fisher’s exact test for dichotomized mRS were used, resulting in Holm-adjusted p-values. A p-value <0.05 was considered statistically significant. The data supporting the findings of this study are available from the corresponding author upon request.

## Results

### Study Cohort and Patient Inclusion

A total of 628 patients with aSAH were screened for eligibility. Sixty-one were excluded due to aSAH-related death within ≤3 days. Of the remaining 567 patients, 166 (29%) developed refractory DCI and underwent a total of 440 IRTs, whereas 401 (71%) did not develop refractory DCI (Figure 1).

### Baseline Demographics and Procedures

Sex distribution differed between groups with a higher proportion of women in the refractory DCI patients (120/166 (72%) vs. 251/401 (63%), p=0.033, Table 1, as for this entire section).

Age also differed (p=0.007) as median age was 56 years (IQR 46–63) vs. 57 years (IQR 48–65) in men, and 55 years (IQR 47–64) vs. 59 years (IQR 50–70) in women with vs. without refractory DCI.

Aneurysm location patterns were comparable between both groups (p=0.870), with anterior circulation sites predominating in both cohorts and similar relative distributions across the major vessels.

Radiographic and clinical aSAH severity differed between groups: Fisher grade 3 and poorer WFNS grades were more frequent in the refractory DCI group (p=0.009 and p=0.012). Assessed only among patients with refractory DCI, BNI score clustered mainly in grades 3–4, and modified WFNS ^61^ most frequently fell into grades 1 and 5. Intracerebral hemorrhage was more frequent in patients with refractory DCI (39/166 [23%] vs. 62/401 [15%]; p=0.030).

Aneurysm treatment modality did not differ between groups (p=0.060). External ventricular drainage was performed more often in refractory DCI (139/166 [84%] vs. 255/401 [64%]; p<0.001), whereas lumbar drainage rates were similar (65/166 [39%] vs. 172/401 [43%]; p=0.454).

### Occurrence, Onset and Duration of refractory DCI

Refractory DCI occurred in 166 (29%) patients (Figure 2). In 14 patients (8.4% of DCI patients, 2.5% of all patients), IRTs for refractory DCI were performed before day 5 after initial aSAH. In 29 patients (17.5% of DCI patients; 5.1% of all patients), IRTs were performed later than day 14 after the aSAH. Median day of first IRT was 8 (IQR 7–11), median day of last IRT was 11 (IQR 9–14), median duration was 2 days (IQR 1–5), and median number of IRTs was 2 (IQR 1–3). No patient aged >70 years needed an IRT later than 14 days after initial aSAH.

**Figure 2.**
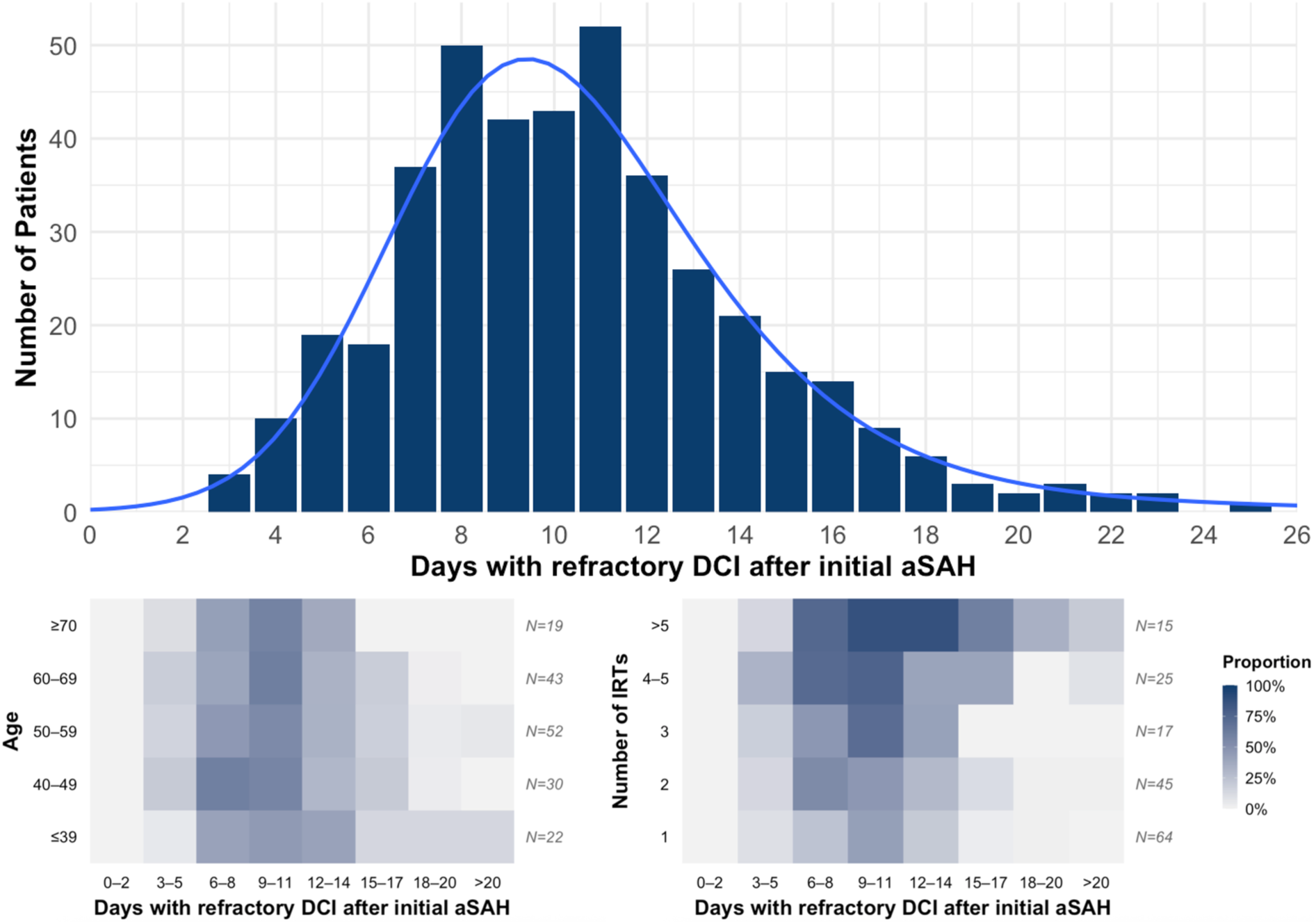
Occurrence of refractory DCI assessed by performed IRTs. Top: All patients with refractory DCI (N = 166 (26.4% of total N = 628)) in relation to the day after initial occurrence of an aSAH. Median day of first DCI was 8 (IQR 7–11), median day of last DCI was 11 (IQR 9–14), median duration was 2 days (IQR 1–5). Bottom left: The same N = 166 broken down by age groups in a heat map. Bottom right: The same N = 166 broken down by number of IRTs in a heat map. DCI = delayed cerebral ischemia, IRTs = interventional rescue therapies, aSAH = aneurysmal subarachnoid hemorrhage

In the multivariable analysis (Table 3), younger age (OR 0.98 per year; p<0.001) and female sex (OR 0.57 for male; p=0.007) were associated with higher odds of developing refractory DCI. A higher WFNS grade was significantly associated with higher odds (OR 1.18 per grade; p=0.011), whereas Fisher grade was not (OR 1.18 per grade; p=0.280).

**Table 3:** Multivariable Models for adjusted Association of multiple Factors regarding Occurrence of Refractory DCI, New Ischemic Stroke at Discharge and Functional Outcome (mRS) in all aSAH Patients (except early deaths ≤3 days)

| Predictor | Effect * | Estimate (95% CI) | p † |
| --- | --- | --- | --- |
| Outcome: Occurrence of Refractory DCI (Multivariable Logistic Regression) |  |  |  |
| Age (per +1 year) | OR | 0.98 (0.96, 0.99) | <0.001 |
| Male sex (vs. female) | OR | 0.57 (0.38, 0.86) | 0.007 |
| Fisher grade (per +1) | OR | 1.18 (0.87, 1.59) | 0.280 |
| WFNS grade (per +1) | OR | 1.18 (1.04, 1.34) | 0.011 |
| Model Summary |  | N = 551, NA = 16, AIC ± = 656.4 |  |
| Outcome: New Ischemic Stroke at Discharge (Multivariable Logistic Regression) |  |  |  |
| Age (per +1 year) | OR | 1.00 (0.98, 1.01) | 0.784 |
| Male sex (vs. female) | OR | 0.89 (0.57, 1.38) | 0.598 |
| Fisher grade (per +1) | OR | 1.40 (0.99, 1.97) | 0.057 |
| WFNS grade (per +1) | OR | 1.27 (1.11, 1.45) | <0.001 |
| Refractory DCI (yes vs. no) | OR | 4.68 (3.06, 7.15) | <0.001 |
| Model Summary |  | N = 518, NA = 49, AIC ± = 561.4 |  |
| Outcome: mRS Score at Follow-Up (3-6 vs. 0-2) (Multivariable Logistic Regression) |  |  |  |
| Age (per +1 year) | OR | 1.06 (1.04, 1.08) | <0.001 |
| Male sex (vs. female) | OR | 1.59 (0.97, 2.59) | 0.067 |
| Fisher grade (per +1) | OR | 1.74 (1.16, 2.61) | 0.008 |
| WFNS grade (per +1) | OR | 1.62 (1.38, 1.89) | <0.001 |
| Refractory DCI (yes vs. no) | OR | 2.37 (1.44, 3.90) | <0.001 |
| Model Summary |  | N = 518, NA = 49, AIC ± = 453.7 |  |

\* Effect (OR): odds ratio for the outcome per 1-unit increase (OR>1 higher odds; OR<1 lower odds)
† Significant p-values are p<0.05 and are shown in bold
‡ AIC: Akaike Information Criterion (lower values indicate better fit, penalized for model complexity)
\* Effect (OR): odds ratio for the outcome per 1-unit increase (OR>1 higher odds; OR<1 lower odds)
† Significant p-values are p<0.05 and are shown in bold

In further multivariable models (Table S3), an earlier first IRT was associated with a longer refractory DCI duration (incidence rate ratio (IRR) 0.88; p<0.001), and a higher number of required IRTs (IRR 0.91; p<0.001). Higher Fisher grade was associated with more IRTs (IRR 1.24; p=0.014), while age, sex, and modified WFNS grade were not significant in these models.

### Impact of DCI: Radiological and Clinical Outcomes

In patients with refractory DCI more ischemic strokes were diagnosed at discharge (91/166 (55%) vs. 73/401 (18%), p<0.001, Table 2). Among patients with new ischemic strokes, frequencies were represented in similar proportions across all IRT categories, including 1, 2, 3, 4–5, and >5 treatments (Figure S2).

In the multivariable model for the occurrence of a new ischemic stroke at discharge, higher WFNS grades (OR 1.27, p<0.001) and refractory DCI (OR 4.68, p<0.001) were significantly associated, whereas higher age (OR 1.00, p=0.784), male sex (OR 0.89, p=0.598), and higher Fisher grade (OR 1.40 per grade, p=0.057) were not (Table 3). Within the refractory DCI cohort, a corresponding multivariable model showed none of the above parameters to reach statistical significance (all p≥0.076) (Table S3).

Median length of stay in tertiary care was longer in patients with refractory DCI (23 days [IQR 18–29] vs. 18 days [IQR 14–22]; p<0.001); results were similar after excluding in-hospital deaths (25 days [IQR 21–31] vs. 19 days [IQR 15–23]; p<0.001) (Table 2). In-hospital mortality was not significantly higher in patients with refractory DCI (27/166 [16%] vs. 48/401 [12%]; p=0.176), whereas 23/27 deaths in the first group were attributed to DCI.

Discharge destination differed between groups (p<0.001), with discharge home being more frequent in patients without refractory DCI (143/401 (36%) vs. 21/166 (13%)) and discharge to rehabilitation or another hospital more frequent in patients with refractory DCI (118/166 (71%) vs. 209/401 (52%)).

Functional outcome at follow-up differed between groups on ordinal comparison (p<0.001) (Table 2, Figure 3). For the dichotomized endpoint, good outcome (mRS 0– 2) was less frequent in patients with refractory DCI (110/159 (69%)) vs. patients without refractory DCI (303/391 (77%), p=0.041, Holm-adjusted p=0.100). Figure 3 summarizes dichotomized follow-up outcomes across subgroups and presents Holm-adjusted comparisons. In the DCI subgroup treated with only 1-2 IRTs (N=103), good outcome was observed in 74% (Holm-adjusted p=0.434), in the subgroup treated with ≥3 IRTs (N=56) good outcome was observed in 61% (Holm-adjusted p=0.036).

**Figure 3.**
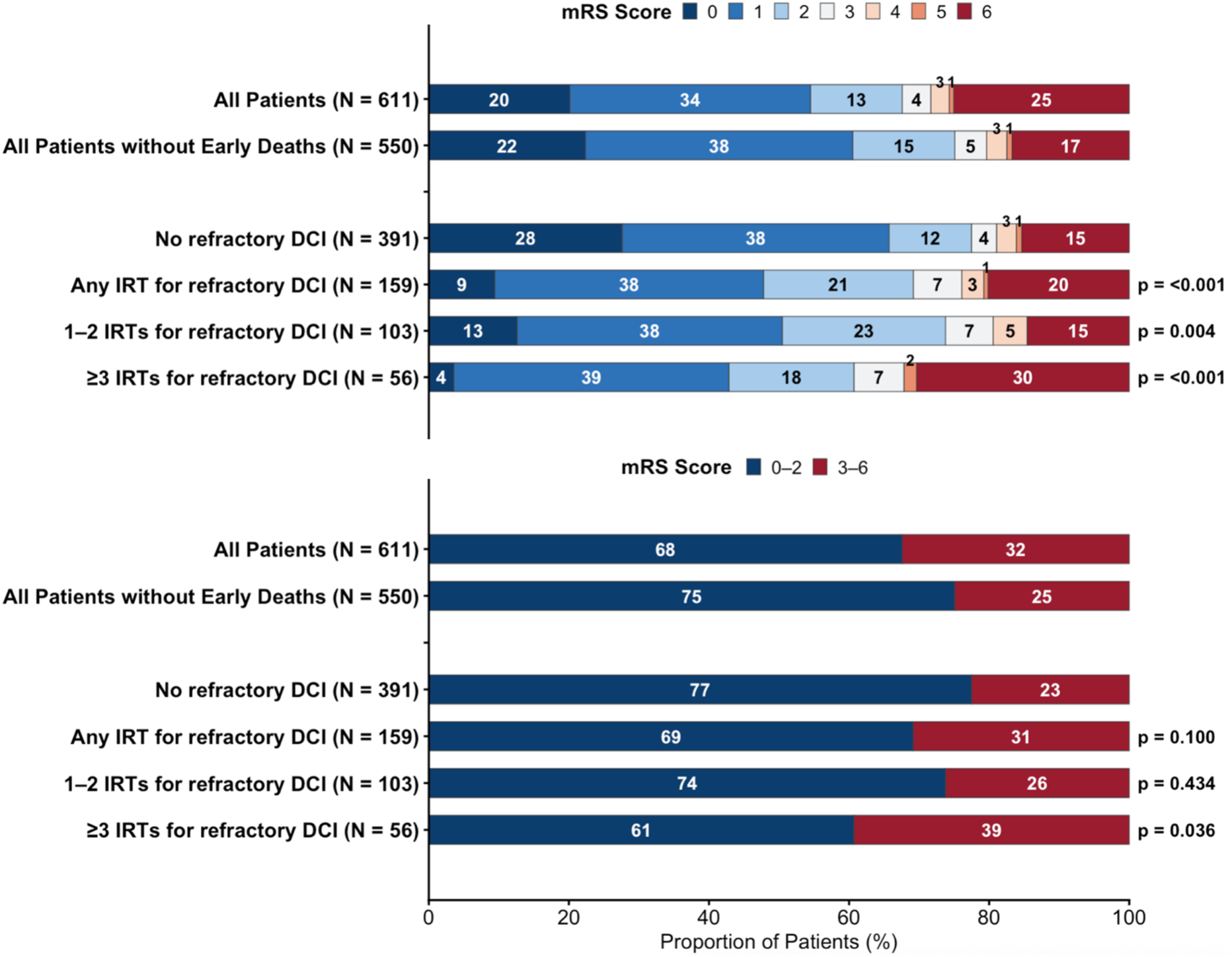
Distribution of mRS at 6 to 12 months across subgroups ‘all patients’ (N = 611 + 17 NAs), ‘all patients except the early deaths ≤ 3 days after initial aSAH’ (N = 550 + 17 NAs) and thereof ‘no refractory DCI’, ‘any IRT’, ‘1–2 IRTs’ and ‘≥3 IRTs for refractory DCI’. Stacked bars show within-group proportions. Right-side annotations show Holm-adjusted p-values for pairwise comparisons versus ‘No refractory DCI’ (Wilcoxon rank-sum shift test for ordinal mRS; Fisher’s exact test for dichotomized mRS). mRS = modified Ranking Scale, aSAH = aneurysmal subarachnoid hemorrhage, DCI = delayed cerebral ischemia, IRT = interventional rescue therapy

In the multivariable model for dichotomized outcome, higher age (OR 1.06, p<0.001), higher Fisher grade (OR 1.74 per grade, p=0.008), higher WFNS grade (OR 1.62 per grade, p<0.001), and refractory DCI (OR 2.37, p<0.001) were significantly associated with poor outcome (mRS 3-6 vs. 0-2, Table 3). Within the refractory DCI cohort, the multivariable model for dichotomized outcome showed higher age (OR 1.04, p=0.019), higher Fisher grade (OR 2.45 per grade, p=0.014), higher modified WFNS grade (OR 1.32 per grade, p=0.028), and earlier day of first IRT (OR 0.86; p=0.026) (Figure S2) to be associated with poor outcome (mRS 3-6 vs. 0-2), whereas male sex was not (OR 1.42, p=0.399) (Table S3).

## Discussion

In this retrospective analysis including more than six hundred patients treated over ten years, refractory DCI affected nearly one third of aSAH patients, occured more frequently in younger women with poorer initial neurological presentation and extended beyond fourteen days in up to a fifth of the patients aged under 70 years. Refractory DCI and earlier DCI onset were associated with a poorer outcome including higher rates of ischemic strokes. In our population of DCI-affected patients, interventional rescue therapies seemed effective in achieving favourable clinical outcomes.

As we applied a strict definition of refractory DCI and only included patients suffering from neurological deficits despite induced hypertension, a direct comparison with previously published work is limited. Nevertheless, the observed DCI incidence of 29% falls within the ranges reported for other aSAH cohorts. ^1,2,5–8^

Regarding risk factors for DCI occurrence, we observed younger age (consistent with findings by four other retrospective analyses ^20,22,23,62^), female sex (consistent with findings by various meta-analyses ^16,17,19,63,64^ and retrospective analyses ^20,35,62^) and poorer initial neurological status (assessed by WFNS score, consistent with findings by three meta-analyses ^16,17,19^ and two retrospective cohorts ^58,65^) associated in multivariable analysis. In accordance with earlier reports, we further observed the amount of subarachnoid blood, assessed by e.g. Fisher scores ^16,17,19,21–23,35,62,66^ to be associated with DCI occurrence.

Timing of DCI onset on median day 8 (7-11) in our patients appeared later compared to three previous analyses with reported onsets between days 3-6, 4-9 and 6-10. ^7,8,67^ This may be explained by the above-mentioned strict inclusion criteria. By comparing the curve of refractory DCI occurrence (Figure 2) with similar curves by Schmidt et al. from 2022 and Tokareva et al. from 2024, a significant difference can be found in the

DCI prevalence > 14 days post ictus. Our prevalence of 17% contrasts with theirs of 1.2% ^8,67^ and 4.4% ^9^ and is supported by assessments of CVS as a radiological correlate of DCI to occur as long as 21 days after aSAH. ^68^ Figure 2 further demonstrates that refractory DCI persisted beyond 14 days only in patients aged < 70. It also shows that in patients requiring more than one IRT, DCI onset was earlier.

Multivariable analyses (Table S3) of risk factors for early / late occurrence and longer duration of DCI as well as its severity (assessed by the number of required IRTs) yielded significant results. Earlier DCI occurrence seemed to be associated with longer duration of DCI and with a higher number of required IRTs. Also younger age, poorer admission WFNS grade and higher Fisher scores appeared to be associated with a higher number of needed IRTs. The only other analysis investigating the timing of DCI that differentiated between an “early” (< 8 days) and “late” (> 8 days) DCI occurrence found younger age and early onset to be significantly associated with longer DCI duration, which aligns with our results. ^8^

New ischemic strokes on imaging at discharge were diagnosed in 18% of non-DCI and 55% of refractory DCI patients. Comparability of these findings remains limited within the available literature, with one recent publication reporting a rate of 13% DCI-related infarctions in a large aSAH population. ^3^ In our cohort, new strokes occurred equally often in patients with one or multiple IRTs and were not found to be associated with a higher number of IRTs (Figure S2). As the details of the ischemic strokes, such as size and pattern, were not assessed, the implications of this finding remain limited. We nevertheless presume that new strokes occurred mainly before the first IRTs, highlighting the importance of close clinical follow-up of patients during the early DCI phase to initiate timely rescue interventions and prevent ischemia.

Regarding risk factors for new ischemic strokes, our results suggest an association with higher Fisher scores and earlier refractory DCI (Figure S2), which aligns with the results by Tokareva et al. ^8^ and Schmidt et al. ^67^ Both found higher rates of strokes in patients suffering from “early” DCI (< day 8 / < day 7). The overall stroke incidence in their DCI cohorts roughly matched our above-mentioned proportion. ^8,67^

Overall clinical outcome was favorable in 77% of patients (mRS of 0-2 at follow-up), which matched the report by La Pira et al. (78%) and was distinctively better than in various other publications (52-64%). ^69–74^ Our multivariable analyses in Tables 3 and S3 reinforce previously suggested risk factors of poor outcome such as higher age ^75–78^, a poorer neurological admission status ^75–78^, greater hemorrhage volumes ^76^ and especially DCI ^3,4,7,78^ as main determinants. Veldemann et al. ^3^ report a 5-fold increase, Raatikainen et al. ^7^ a 2.7-fold increase in odds of poor outcome at one year due to DCI after aSAH, which exceeds our finding of a 2.4-fold increase. In addition, our analysis identifies early occurrence of refractory DCI as a significant predictor of poor outcome. However, and importantly, refractory DCI was not associated with worse outcome in general (Figure 3): While the Fisher’s exact test (Table 2) suggests a significant difference in dichotomized outcome, the more refined Holm-adjusted p-values only reach significance for patients who required three or more IRTs for severe refractory DCI. Hence, patients with refractory DCI who received one or two IRTs had outcomes comparable to those of patients who did not need IRT. This finding provides evidence for the efficacy of IRTs in refractory DCI. The only randomized controlled trial evaluating rescue therapies had previously been stopped early due to increased rates of serious complications of endovascular treatment. ^44^

### Limitations

This study has several limitations, which can be mainly accorded to the retrospective and single-center design of the study. While our findings provide evidence supporting the use of IRTs for DCI, causal conclusions regarding the efficacy of IRTs cannot be made in the absence of a control group. Treatment allocation was clinically driven, resulting in likely selection bias. Further, the present study does not allow for a detailed evaluation of individual IRT modalities. This limitation is partly attributable to the heterogeneity of IRTs performed within our cohort. Although all patients with DCI were primarily treated with intra-arterial bolus nimodipine administration, 37 patients underwent additional balloon angioplasty, 15 received intrathecal nimodipine, and eight were additionally treated with continuous intra-arterial nimodipine.

### Conclusions

This study contributes to the understanding of refractory DCI, particularly affecting younger women with poor neurological status at admission, as an important determinant of outcome after aSAH.

As earlier onset of refractory DCI appears to be associated with worse outcome, and a relevant proportion of patients younger than 70 years experience refractory DCI beyond 14 days after ictus, we recommend early and vigilant neuro-intensive care monitoring, with extension beyond the conventional observation period when clinically indicated. The absence of a detected outcome difference after 1–2 IRTs suggests that favorable outcomes may remain achievable despite refractory DCI.

## Data Availability

All data referred to in the manuscript are available on request.

## Non-standard Abbreviations and Acronyms

aSAH: aneurysmal Subarachnoid Hemorrhage
CVS: Cerebral Vasospasm
DCI: Delayed Cerebral Ischaemia
DSA: Digital Subtraction Angiography
EVD: External Ventricular Drainage
HRA: Human Research Act
HRO: Human Research Ordination IRR Incidence Rate Ratio
IRT: Interventional Rescue Therapy
mRS: modified Ranking Scale

## Acknowledgments

None.

## Supplemental Material

1. Supplemental Figures and Figure Legends

**Figure S1.** Heat maps demonstrating the association between the occurrence of a new ischemic stroke at discharge and the number of required IRTs for refractory DCI (top left), the days of refractory DCI after initial aSAH (top right) and the mRS at follow-up (bottom left) as well as the association between the functional outcome (mRS at follow-up) and the number of required IRTs for refractory DCI (middle left), the days with refractory DCI after initial aSAH (middle right) and the day of first performed IRT (bottom right). All analyses on the left were performed on the N = 567 (NA = 17) patients with an aSAH excluding the deaths within 3 days, all analyses on the right only on the N = 166 (NA = 7). IRT = interventional rescue therapy, DCI = delayed cerebral ischemia, aSAH = aneurysmal subarachnoid hemorrhage, mRS = modified Ranking Scale

2. Supplemental Tables

**Table S1.** Collinearity Assessments by Spearman correlation matrix and VIF/GVIF-equivalents

**Table S2.** Subset Table for Demographics, Characteristics, and Procedures – including only the patients who died within 3 days after aSAH (N=61)

**Table S3.** Multivariable Models in N = 166 Patients interventionally treated for refractory DCI

